# Longitudinal changes in motor network activity and connectivity in patients with suspected Transcient Ischemic Attack: an exploratory fMRI study

**DOI:** 10.64898/2026.10.13.26365278

**Authors:** Guillaume Charbonnier, Louise Bonnet, Alexandre Comte, Elisabeth Medeiros de Bustos, Mathieu Lesourd

## Abstract

**Background and Objectives:** Transient ischemic attack (TIA) diagnosis is challenging because it relies mainly on clinical assessment and exclusion of common differential. Functional MRI (fMRI) may provide a diagnostic biomarker by detecting transient brain dysfunction not visible on conventional imaging. This pilot study aimed to characterize acute motor-task BOLD activation in patients with suspected DWI-negative TIA and compare functional patterns between confirmed TIA and alternative diagnoses. Secondary objectives were to assess longitudinal changes in activation and connectivity at 3 months.

**Methods:** In this prospective single-center study, adults presenting within 24 h of transient anterior-circulation neurological symptoms underwent conventional MRI and additional motor-task fMRI. Patients with acute DWI lesions were excluded. Final diagnosis was established after 3-month follow-up. Whole-brain analyses, individualized motor cortex region-of-interest (ROI) analyses, and generalized psychophysiological interaction (gPPI) connectivity analyses were performed during the acute phase and at 3 months.

**Results:** Eight patients were enrolled; six had complete longitudinal imaging data and were included in the final analyses. Final diagnoses were TIA (n=2), migraine aura (n=2), functional neurological disorder (n=1), and hypoglycemia (n=1). Whole-brain and ROI analyses showed reduced bilateral motor-related BOLD responses during the acute phase in both TIA patients and in the patient with hypoglycemia. In contrast, patients with migraine and functional neurological disorder exhibited less marked longitudinal changes. In TIA patients, motor cortex activation increased at 3 months, tending to normalize toward values observed in non-TIA participants. Connectivity analyses revealed heterogeneous patterns, but the 2 TIA patients demonstrated increased longitudinal connectivity between the motor cortex contralateral to symptoms and other sensorimotor regions, including supplementary motor, premotor, parietal, and postcentral cortices.

**Discussion:** These preliminary findings suggest that TIA may be associated with transient bilateral reductions in motor-network activity. Functional abnormalities appeared to improve over time, supporting the hypothesis of reversible network dysfunction. Although limited by the very small sample size, this study provides the first longitudinal assessment of emergency task-based fMRI in rigorously defined TIA and supports further investigation of acute functional MRI as a potential biomarker of transient cerebral dysfunction.

## Introduction

Transient ischemic attack (TIA) was traditionally defined as a transient neurological deficit resolving within 24 hours. Advances in neuroimaging, particularly the use of diffusion-weighted MRI (DWI), have led to a tissue-based definition of TIA as “a transient episode of neurological dysfunction caused by focal brain, spinal cord, or retinal ischemia, without acute infarction”^1^. Prompt diagnosis and management, including identification of the underlying etiology and implementation of secondary preventive measures, are essential to reduce the risk of recurrent transient ischemic attacks or ischemic strokes. TIA diagnosis consequently relies on specialized neurological assessment, based on patient history, and combined with appropriate MRI sequences, particularly DWI during the acute phase, to exclude a clinically reversible infarction. However, acute-phase MRI is not routinely available worldwide, limiting the extent to which this tissue-based definition can be applied in clinical practice and in research. Clinically, sudden-onset unilateral motor symptoms lasting more than 2 minutes are generally considered to provide a high level of suspicion for TIA. Nevertheless, even these presentations may result from alternative diagnoses, most notably migraine aura, epileptic seizures, and functional neurological disorders. Improving our understanding of the pathophysiological mechanisms underlying TIA and developing more reliable diagnostic approaches are therefore crucial. Beyond the clinical diagnosis itself, an important question is whether the apparent resolution of symptoms reflects a complete normalization of brain function. The absence of a detectable acute lesion on conventional neuroimaging does not necessarily imply that brain structure and function have returned to their pre-event state. Recent findings suggest that clinical recovery does not necessarily reflect a full return to the pre-morbid neurofunctional state. The condition may therefore be associated with persistent structural^1^ and functional brain alterations^2^, despite the apparent resolution of clinical symptoms. However, there is currently no similar study with a strict TIA definition using DWI-negative, less than 24h after symptoms onset MRI. Moreover, there is no longitudinal data on acute functional abnormalities.

The aim of this preliminary study is to describe BOLD functional activation at the acute phase of TIA suspected patients and compare TIA final diagnosis with differential diagnosis. Secondary objective is to compare 24h functional abnormalities with a 3-month follow up MRI scan.

## Methods

### Participants

We conducted a prospective, single-center, exploratory, comparative, interventional study between October 2019 and April 2021. The study was approved by the Ethics Committee (CPP) on November 20, 2018, reference 2018-A00762-53. All patients signed a written informed consent form before being included in the study. Inclusion criteria included any patient over 18 years of age presenting with a sudden transient neurological deficit suggestive of an anterior circulation TIA within 24 hours: partial or complete sensory or motor disturbance of one side of the body, partial or complete aphasia, with a prescription for conventional MRI within 24 hours of symptom onset, including at least an urgent DWI sequence to search for an ischemic lesion. Patients with DWI lesions, a history of neurological lesions, severe psychiatric illness (current major depressive episode, schizophrenia, bipolar disorder), or brain pathology discovered during the initial MRI were excluded. If patients participated in the study, the MRI was supplemented with three research sequences: right and left motor activation fMRI, and resting state fMRI. These sequences are detailed below. A standardized assessment was then performed as part of routine care to diagnose TIA: conventional MRI (DWI, FLAIR T2*, ToF) within 24 hours of symptom onset, Doppler ultrasound of the supra-aortic trunks, blood tests (complete blood count, lipid profile, glycated hemoglobin, TSH), to rule out a differential diagnosis (electroencephalogram), to screen for an embolic cardiac arrhythmia (telemetry), and to screen for cognitive impairment (Montreal Cognitive Assessment (MoCA), version 1). Patients were then systematically reviewed at 3 months with a neuropsychological assessment, screening for mood disorders, a neurological examination (departmental protocol), and a follow-up brain MRI including functional MRI sequences. It is during this visit that the diagnosis of TIA is confirmed or ruled out, and that patients are classified into the TIA or non-TIA group by a neurologist specializing in all clinical and paraclinical data since the onset of the regressive deficit. The neuropsychological assessment lasts approximately 45 minutes and includes: the Montreal Cognitive Assessment version 3 (MoCA^3^); the Stroop test (Stroop victoria version^4^ or DKEFS version^5^); the Trail Making Test (DKEFS version^5^); the Rey-Osterrieth Complex Figure Test^6^), the Oral Naming Test (DO30^7^); auditory-verbal working memory, short memory, and coding subtests (WAIS IV^8^), Fluency (‘Animals’ and ‘P’^9^), cancellation letter task (i.e., letter A^10^); Phasic alertness (TAP^11^), visual episodic memory ( Rey-Osterrieth Complex Figure) and verbal episodic memory (RL/RI-16^12^ or CVLT^13^ or TMA-93^14^). The screening for psychiatric disorders includes: the Hamilton Anxiety and Depression Scales and the Multidimensional Fatigue Inventory.

### fMRI

fMRI scans were performed at two time points: during the acute phase (<24 h after motor symptoms onset) and during the chronic phase (about 3 months post episode).

#### Procedure and design

A block design consisting of 4 alternating movement and rest epochs was employed. Each movement block and each rest block lasted 15 s. Functional MRI acquisition comprised two separate runs. During one run, participants performed repeated movements with the right hand only, whereas during the other run they performed the same task with the left hand only. The right-hand movement run was always performed first, followed by the left-hand movement run.

#### fMRI data acquisition

Imaging data were acquired on a 3T Siemens Vida XT Scanner (Siemens, Erlangen, Germany) using a 64-channel head coil. Blood-Oxygen Level Dependent (BOLD) fMRI data were acquired using a T2*-weighted echo-planar imaging (EPI) sequence with Simultaneous Multi-Slice (SMS) acceleration (multiband factor = 3) at both acute and chronic phases. Functional images were all collected as oblique-axial scans aligned with the anterior commissure–posterior commissure (AC–PC) line with the following parameters: 135 volumes per run (270 volumes at each phase), 42 slices, TR/TE = 1000 ms / 30 ms, flip angle = 90°, field of view = 234 x 234 mm^2^, slice thickness = 3 mm, voxel size = 2.7 x 2.7 x 3 mm^3^. Structural T1-weighted images were collected using an MPRAGE sequence (176 sagittal slices, TR/TE = 1700 / 2.55 ms, inversion time = 900 ms, flip angle = 9°, 240 x 256 mm FOV, slice thickness = 1 mm, voxel size = 0.5 x 0.5 x 1 mm^3^) only during the chronic phase.

#### Preprocessing of fMRI data

Structural T1-weighted images were segmented into tissue type (GM: grey matter, WM: white matter and CSF: cerebro-spinal fluid tissues) using the Computational Anatomy Toolbox (CAT12; http://dbm.neuro.uni-jena.de/cat12/) segmentation tool, in order to facilitate the normalization step. Functional data were analyzed using SPM12 (Wellcome Department of Cognitive Neurology, http://www.fil.ion.ucl.ac.uk/spm) implemented in MATLAB (Mathworks, Sherborn, MA). Preprocessing for univariate analyses included the following steps (1) 3D motion correction and linear detrending; (2) realignment to the mean EPI image with 6-head motion correction parameters and unwarping using topup (Andersson et al. 2003); (3) co-registration of the individual functional and anatomical images; (4) normalization towards MNI template; and (5) spatial smoothing of functional images (Gaussian kernel with 5 mm FWHM).

#### GLM

A general linear model was created using design matrices containing one regressor for each movement condition (left-hand movement and right-hand movement) modeled as a boxcar function (with onsets and durations corresponding to the start of each movement condition) convolved with the canonical hemodynamic response function (HRF). Six regressors of non-interest resulting from 3D head motion estimation (x, y, z translation and three axis of rotation) were added in the design matrix. The model was estimated in each patient, also taking into account the average signal in the run. After model estimation, we computed first-level contrasts against baseline for each hand movement (i.e., right- and left-hand movements) at T0 (acute stage) and T1 (chronic stage). To assess longitudinal changes, we also computed within-subject contrasts comparing the chronic and acute sessions (T1 > T0) separately for right- and left-hand movements. To assess longitudinal changes, we computed within-subject contrasts comparing T1 > T0 separately for right- and left-hand movements. As we considered these patients as a case series, we did not transfer to a second level group analysis. Moreover, to further characterize motor-related activation patterns, ROI analyses were performed (see the ROIs selection section). Results maps are presented with a significance threshold set at *p* < .05 with family-wise error (FWE) correction for simple contrasts with an additional conservative threshold (*k* = 100 voxels).

#### ROIs selection

For each patient, the left and right M1 regions were functionally defined using a multi-step procedure (see **supplementary Figure 1**^15^). First, Neurosynth was used to identify brain regions consistently associated with hand-related tasks. The terms “*hand*” was entered into the Neurosynth database to generate a meta-analytic functional activation map reflecting the brain regions consistently activated across studies using this term^1^ (FDR corrected, *p* < .01). This map was used to identify the core motor network involved in hand movements and to provide an initial functional constraint for the subsequent patient-specific functional localization of M1. The clusters corresponding to the motor cortex were selected in each hemisphere. Second, for each patient, we identified within each functional cluster the voxel showing the highest *t*-value in the GLM contrast “*all conditions vs. baseline*”. This contrast was used to identify patient-specific task-responsive voxels. Third, a 6-mm-radius sphere was then defined around each peak voxel, yielding two individual ROIs per patient (see **supplementary Figure 1** and **supplementary Table 1**). This patient-specific approach accommodated interindividual variability in the precise location of peak activity while ensuring that all individual peaks remained contained within the functional cluster map.

To obtain a single beta estimate per condition and per patient for each ROI, we used the MarsBaR toolbox v0.45 (Brett et al., 2002; https://marsbar-toolbox.github.io) in Matlab. MarsBaR computes a separate GLM for each ROI and patient using the original first-level design matrix and contrast definitions. This procedure yielded one beta value per condition and per patient in each ROI. Parameter estimates (β values) were extracted from the two individually-defined spherical ROIs described above, for left and right-hand movements.

#### Functional connectivity analyses

Analyses of fMRI data were performed using CONN^16^ (RRID:SCR_009550) release 22.v2407^17^ and SPM^18^ (RRID:SCR_007037) release 12.7771.

#### Denoising

In addition, functional data were denoised using a standard denoising pipeline^[4]^ including the regression of potential confounding effects characterized by white matter timeseries (5 CompCor noise components), CSF timeseries (5 CompCor noise components), SPM_covariates regressors (6 components), and linear trends (2 factors) within each functional run, followed by bandpass frequency filtering of the BOLD timeseries^19^ between 0.008 Hz and 0.09 Hz. CompCor^20,21^ noise components within white matter and CSF were estimated by computing the average BOLD signal as well as the largest principal components orthogonal to the BOLD average within each subject’s eroded segmentation masks. From the number of noise terms included in this denoising strategy, the effective degrees of freedom of the BOLD signal after denoising were estimated to 76.8 across all subjects^22^.

#### General Psychophysiological interactions (gPPI)

Psychophysiological interaction analyses were used to investigate task-dependent changes in functional connectivity. Seed regions included 10 predefined ROIs: individually defined spherical ROIs in the left and right primary motor cortex (M1), together with bilateral precentral (PreC) gyrus, superior parietal lobule (SPL), postcentral (PostC) gyrus, and supplementary motor area (SMA) ROIs derived from the CONN atlas. Separately for each pair of seed and target areas, a generalized psychophysiological interaction model (gPPI^23,24^) was defined with seed BOLD signals as physiological factors, boxcar signals characterizing each individual task condition convolved with an SPM canonical hemodynamic response function as psychological factors, and the product of the two as psychophysiological interaction terms. Functional connectivity changes across conditions were characterized by the Fisher-transformed semipartial correlation coefficient of the psychophysiological interaction terms in each model. Although gPPI analyses were computed across all pairs of predefined sensorimotor ROIs, we focused our interpretation on task-dependent connectivity changes involving the left and right primary motor cortices (M1) and the remaining motor-related regions. No second-level group analysis was performed due to the limited sample size. Fisher-transformed gPPI connectivity values were therefore inspected at the individual level to characterize patterns of task-dependent network reorganization.

## Results

Eight patients were included in the study, which had to be prematurely stopped due to technical incidents and the COVID-19 pandemic, which prevented recruitment. Due to missing data at either the acute or chronic phase, complete analyses could not be performed for patients 3 and 6^2^; consequently, they are not included in the analyses presented below.

### Demographic and clinical data

Patients’ characteristics are presented in Table 1. Their ages ranged from 28 to 68 years, and the sample included three males and three females. All patients were right-handed, although handedness data were missing for patient 3.

**Table 1.** Demographic and clinical data of the patients.

| Patient n° | Age | Sex | Handedness | IRM delay (hh:mn) | symptom type (motor/sens/speech/head./vis.) | symptom side | symptom duration (hh:mn) | Clinical diagnosis |
| --- | --- | --- | --- | --- | --- | --- | --- | --- |
| 1 | 20-30 | F | Right | 5:20 | 1/1/0/1/0 | left | 01:00 | Migraine |
| 2 | 20-30 | F | Right | 15:23 | 0/1/0/0/0 | Right | 01:00 | Migraine |
| 3 | 40-50 | M | NA | 10:29 | 1/1/0/1/1 | Right | 01:00 | Migraine |
| 4 | 30-40 | M | Right | 9:12 | 1/1/0/1/0 | Right | 00:30 | TIA |
| 5 | 40-50 | M | Right | 7:10 | 1/1/1/0/0 | Left | 01:00 | Hypoglycemia |
| 6 | 60-70 | F | Right | 4:22 | 1/0/0/0/0 | Left | 00:30 | TIA |
| 7 | 50-60 | F | Right | 11:33 | 1/0/1/0/0 | Left | 00:45 | FND |
| 8 | 50-60 | F | Right | 4:19 | 1/0/0/0/0 | Right | 00:15 | TIA |

sens.: sensitive; head.: headache; vis.: visual; TIA: transient ischemic attack; FND Functional Neurological Disorder; NA: Non-Available data.

### Neurosychological data

Neuropsychological data are presented in **supplementary Table 1**. An initial MoCA score was available for P1, P2, P5, and P6 (range: 26–29). Among patients with MoCA scores available at 3 months post-episode, scores remained relatively stable, except for P6, who showed a 3-point decrease. A neuropsychological assessment was performed for all patients at 3 months, except P3. Overall, no evidence of global cognitive dysfunction was observed, except in P7, who had a lower MoCA score and showed attentional deficits and reduced processing speed. TIA patients (P4 and P8) showed preserved cognitive functioning at the 3-month assessment.

### fMRI univariate results

#### Whole-brain analysis

Whole-brain contrasts of left- and right-hand movements against baseline at T0 and T1 are presented in **Figure 1**. According to these analyses, the two TIA patients (P4 and P8), as well as the patient with hypoglycemia (P5), tended to show lower BOLD responses during the acute phase compared with the chronic phase, in contrast to the less pronounced longitudinal changes observed in the other patients. Although this pattern appeared consistent when examining the longitudinal contrast (T1 > T0) in these three patients, increased BOLD responses at T1 compared with T0 were also observed in the patient with functional neurological disorder (P7) and in patients with migraine aura (P1 and P2).

**Figure 1.**
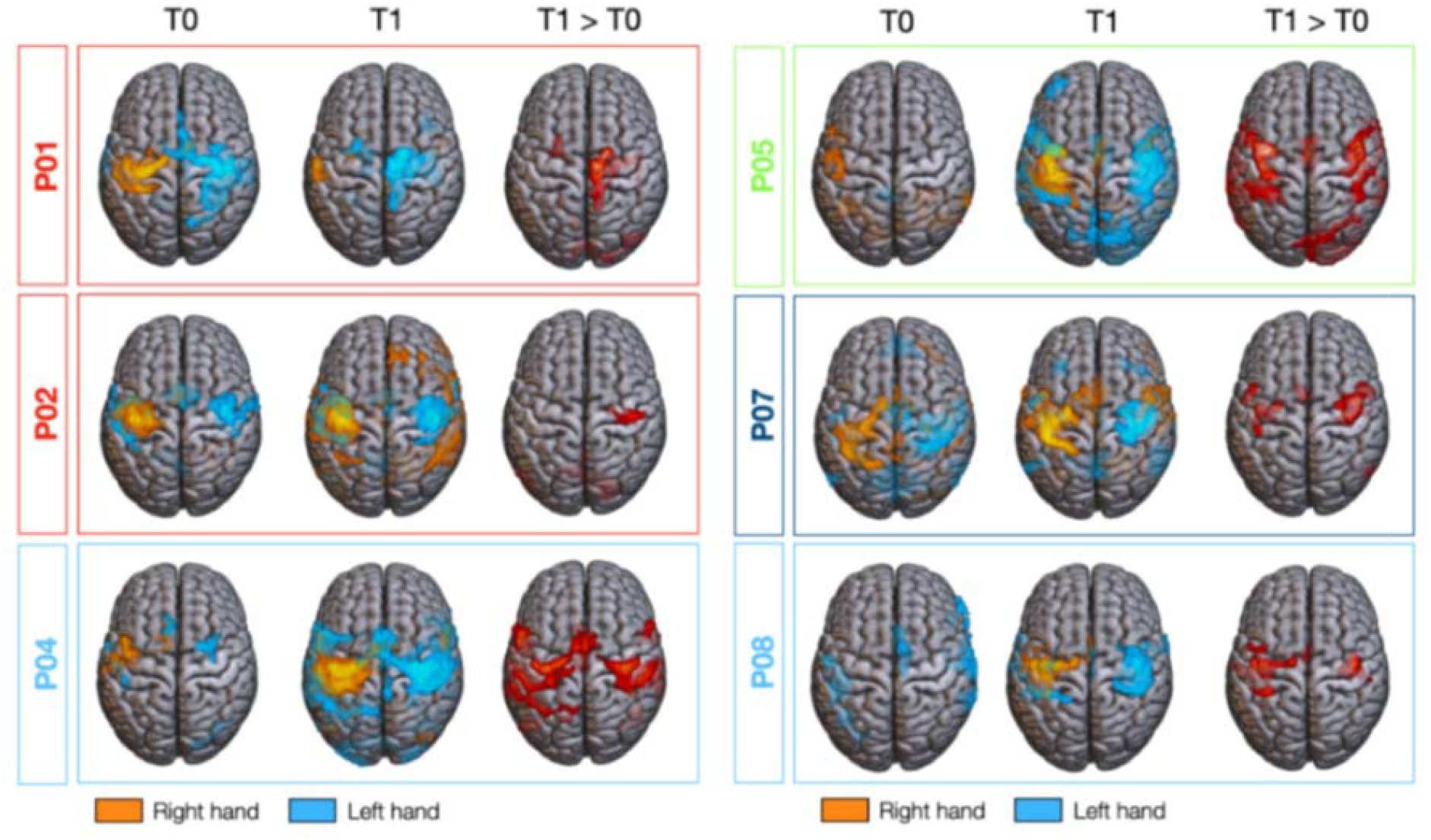
Brain activation during left- and right-hand movement tasks at the acute (T0) and chronic (T1) phases following the episode. Direct comparisons between T1 and T0 are also presented for each patient. Statistical maps are FWE-corrected (*p* < .05) with an extent threshold of 100 voxels, and displayed on an MNI152 template. Patients are color-coded according to their final diagnosis: transient ischemic attack (TIA, light blue), migraine (red), hypoglycaemia (green) and functional neurological disorder (FND, dark blue).

#### ROI analysis

To further characterize longitudinal changes in BOLD signal between T0 and T1, we performed ROI analyses centered on individually defined functional motor regions in the left and right hemispheres, separately for each contralateral hand movement. ROI analyses further supported the pattern observed at the whole-brain level, with patients with TIA, together with the patient with hypoglycemia, showing numerically lower β values at T0 compared with the other patients. At T1, β values displayed greater inter-individual variability (see **Figure 2** and supplementary **Table 3**). In contrast, P1 exhibited increased BOLD responses during the acute phase, potentially related to the persistence of migraine aura symptoms at the time of scanning (about 5 hours delay from symptom onset). Importantly, TIA patients tended to show an increase in motor-related activation from T0 to T1, with beta values becoming more similar to those observed in the other patients at T1. This pattern may support the potential reversibility of the acute-phase alterations in motor brain regions, or at least a partial normalization of motor-related activity over time.

**Figure 2.**
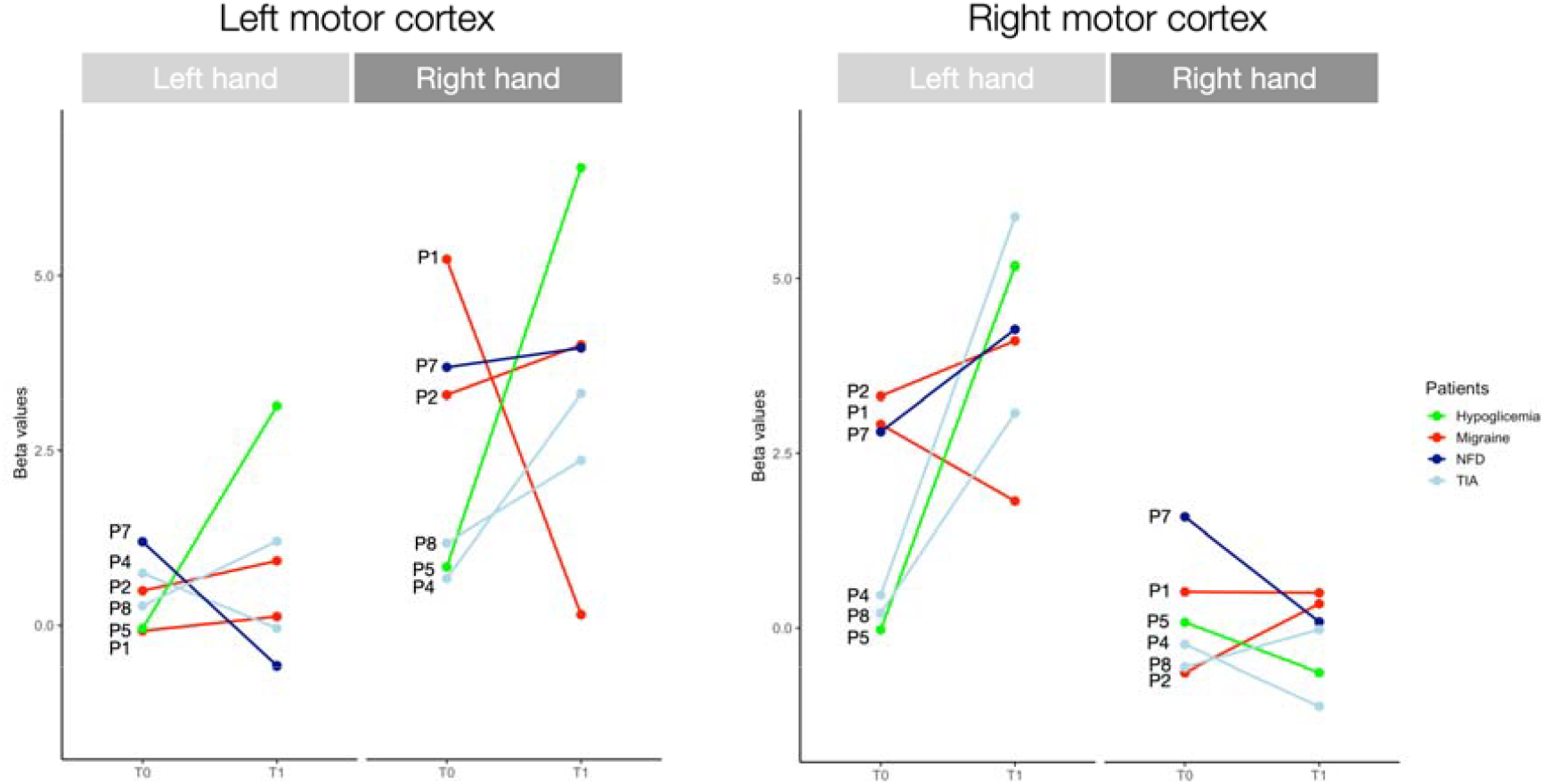
Individual beta values extracted from the left and right sensorimotor cortex during left- and right-hand motor tasks in the acute phase (T0) and at 3-month follow-up (T1). Each line represents one patient. Patients are color-coded according to their final diagnosis: transient ischemic attack (TIA, light blue), migraine (red), hypoglycaemia (green) and functional neurological disorder (FND, dark blue). The left panel shows beta values extracted from the left motor cortex, and the right panel shows beta values extracted from the right motor cortex. Separate facets correspond to left-hand and right-hand motor execution conditions.

No clear association was observed between symptom laterality and beta values in the TIA patients. Although both patients presented with right-sided motor and/or sensory symptoms, beta values were relatively close between the left and right motor cortices (P4 left M1 for right-hand movement = 0.67 and right M1 for left-hand movement = 0.47; P8 left M1 for right-hand movement = 1.18 and right M1 for left-hand movement = 0.21). Thus, the decrease in beta values did not appear to be specifically associated with the motor cortex contralateral to the symptomatic side. Rather, the similar beta values observed across the two motor cortices may suggest a more bilateral decrease in motor cortex activation, although this interpretation remains preliminary and should be further investigated in a larger sample. Interestingly, TIA patients tended to show a larger increase in beta values from T0 to T1 in the motor cortex contralateral to the side of their initial symptoms. Both patients with right-sided symptoms showed a greater increase in beta values in the left M1 than in the right M1 (P4: Δβ = 5.40 vs. 2.64; P8: Δβ = 2.87 vs. 1.18, respectively). This pattern may suggest that, although both hemispheres may be affected during the acute phase of TIA, the motor cortex contralateral to the symptomatic side may show a greater recovery between the acute and follow-up stages.

### Longitudinal changes in functional connectivity (gPPI)

The longitudinal analysis of changes in functional connectivity (ΔFC = T1−T0) revealed heterogeneous individual profiles, whose organization appeared to depend on both clinical lateralization and the side of the hand being moved (see **Figure 3**). In patients presenting with right-sided symptoms (P2, P4, and P8), the left M1, i.e. the M1 contralateral to the symptoms, generally showed positive changes in connectivity with several regions of the motor network, including the premotor and postcentral cortices, SMA, and SPL. This pattern was particularly evident in the two patients who had experienced a TIA (P4 and P8), although their profiles differed substantially in terms of their dependence on the side of the hand being moved.

**Figure 3.**
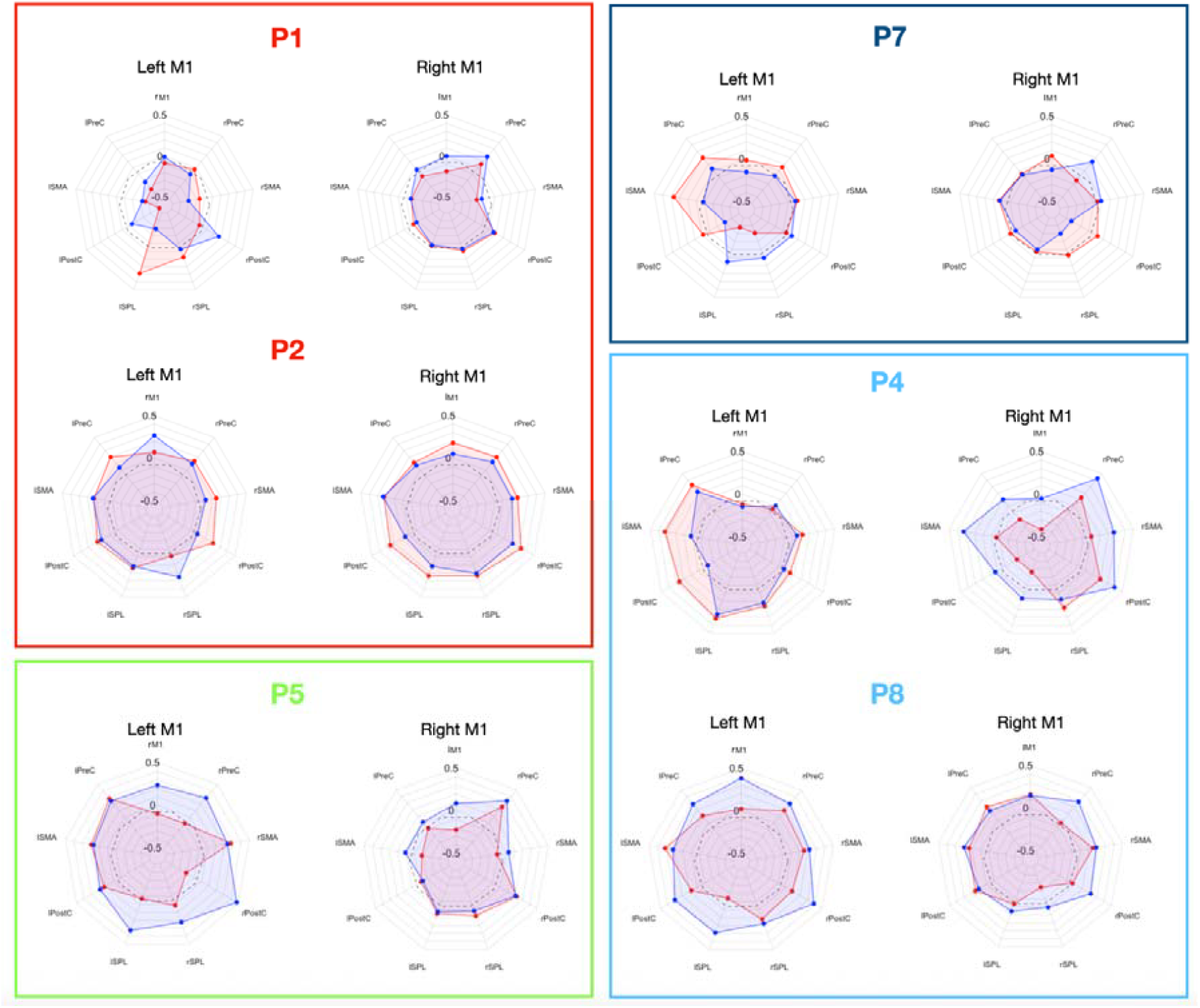
Longitudinal changes (T1–T0) in functional connectivity between M1 cortices and sensorimotor regions during left- and right-hand movements. Values represent the difference in Fisher z-transformed connectivity scores between T1 and T0 (ΔFC = T1 − T0). Patients are color-coded according to their final diagnosis: transient ischemic attack (TIA, light blue), migraine (red), hypoglycaemia (green) and functional neurological disorder (FND, dark blue). M1: primary motor individual ROI; PreC: precentral gyrus; SMA: supplementary motor area; PostC: post-central gyrus; SPL: superior parietal lobe; l/r: right/left.

In P4, increases in connectivity within the left M1 were particularly pronounced during movement of the right symptomatic hand. In this condition, left M1 showed increased connectivity with the premotor cortex (+0.32), SMA (+0.33), postcentral cortex (+0.26), and SPL (+0.32). These changes were generally larger than those observed during movement of the left, non-symptomatic hand. P4 therefore displayed a profile in which longitudinal changes in the contralateral motor network appeared relatively specific to recruitment of the symptomatic limb. However, this increase was not generalized across the entire network, as some connections involving the right M1, particularly with several regions in the left hemisphere, decreased concurrently. The profile of P8, who also had a TIA with right-sided symptoms, was substantially different. Although increased connectivity of the left M1 was observed during movement of the right symptomatic hand, positive changes were even more widespread during movement of the left, non-symptomatic hand. In this condition, left M1 showed increased connectivity with the contralateral M1, premotor cortex, SMA, postcentral cortex, and SPL, as well as with several homologous regions in the right hemisphere. The profile of P8 therefore appeared more diffuse and bilateral, with no clear specificity for recruitment of the symptomatic limb.

The other patients showed more heterogeneous patterns of longitudinal connectivity change. P2, who also presented with right-sided symptoms, showed a more moderate increase in connectivity of the contralateral M1 under both motor conditions, with only a slight predominance during movement of the symptomatic hand. Among patients with left-sided symptoms, P5 showed a pattern qualitatively similar to P4, with greater increases in contralateral M1 connectivity during movement of the symptomatic hand, whereas P1 and P7 showed weaker or predominantly negative changes. Overall, these additional profiles indicate that increases in motor-network connectivity are not specific to TIA, nor are they systematically associated with movement of the symptomatic limb. Rather, they appear to reflect heterogeneous individual changes in the functional organization of the motor network.

## Discussion

We highlighted a novel MRI biomarker for TIA, with an original pilot study using 1-a DWI-negative definition of TIA, 2-a 24h emergency fMRI scan, 3-a longitudinal fMRI control at 3 months. Among 6 patients included in the final analysis, 2 patients were diagnosed with TIA, 3 patients with a differential diagnosis (2 migraines and 1 FND), and 1 was after the fact, ineligible with a final diagnosis of hypoglycemia. Initial fMRI scans demonstrated 4 patients with Bold anomalies compared to the same patients at 3 months (whole brain analysis and GPPI). the 2 patients with TIA and the patient with hypoglycemia demonstrated bilateral hypoactivities in Bold fMRI with motor tasks. The 3 other patients did not show differences at the 2 time points. The shared anomalies between TIAs and hypoglycemia highlight the common hypothesis that TIA induces a hypometabolism due to diminution of cerebral blood flow. The bilateral anomalies in TIAs are more surprising since arterial stroke concerns mainly one vascular territory but is consistent with previous works. Pivotal studies have described changes in motor activity after strokes, in both hemispheres^25^. Li et al. performed resting state fMRI in 21 TIA and 21 HC, and found decreased connectivity in multiple networks, on both sides^26^. Hao et al. described 48 patients with TIA with MRI scans performed 6h to 16 days after symptoms^27^. Compared to 41 healthy controls they found bilateral M1 and multiple alteration of brain connectivity. Lv et al. performed resting state fMRI in 51 TIA and 41 healthy controls and found decreased Amplitude of low frequency fluctuation (ALFF) in the left middle temporal gyrus and a decreased degree centrality (DC) in the right inferior frontal gyrus in TIA patients^28^.

However, these previous works did not use a strict definition of TIA. To explain these results, we could hypothesize that even a focal, minor ischemia could induce a larger functional alteration, including contralateral network disorganization. This contralateral decreased cortical activity could also be a factor linked to the excellent clinical recovery of TIAs. It has been described that in stroke contralateral hyperactivity is linked to worse recovery, and is a potential target for transcranial brain stimulation. Indeed cathodal stimulation of contralateral hemisphere has been associated with good recovery in AIS^29^.

Resting-state fMRI studies have further provided evidence of persistent alterations in large-scale brain network organization following TIA. At the whole-brain level, Lv et al. reported decreased global network efficiency, reduced nodal centrality in the bilateral insula and basal ganglia, and impaired interhemispheric connectivity in TIA patients assessed between 6 hours and 16 days after the event^28^. These findings suggest that TIA may transiently disrupt the integration and communication of distributed brain networks, beyond the absence of detectable structural lesions. Alterations in resting-state functional connectivity have also been reported at the level of specific resting-state networks (RSNs). Within one month following TIA, patients exhibited both increased and decreased functional connectivity within the default mode network (DMN) and self-referential network (SRN), as well as decreased connectivity within the dorsal attention network (DAN), central executive network (CEN), core network (CN), somatomotor network (SMN), visual network (VN), and auditory network (AN)^26^. Notably, this study did not identify significant associations between functional connectivity within these networks and neuropsychological performance. Moreover, MoCA scores did not significantly differ between TIA patients and controls and were not correlated with RSN connectivity measures. Changes in task-residual effective connectivity during fist-closing movements have also been reported in patients with TIA^2^. Compared with healthy controls, TIA patients showed significantly stronger effective connectivity toward the ipsilateral primary motor cortex (M1), consistent with previous findings in stroke patients showing reduced inhibition and/or stronger excitatory influences toward ipsilateral M1 during movements of the paretic hand^30^. TIA patients also exhibited lower effective connectivity toward the supplementary motor area (SMA) than healthy controls. These results imply that TIA patients have aberrant connections among motor regions, and these changes may reflect the decreased efficiency of primary motor function. Importantly, the TIA population in Li et al.^1,26^ and Chu et al^2^ were not specifically defined by the absence of an acute diffusion-weighted imaging (DWI) lesion. Therefore, these findings cannot be considered evidence that functional connectivity alterations persist specifically in tissue-negative TIA. Nevertheless, taken together with the structural and effective connectivity findings described above, they suggest that an apparently transient TIA may be associated with alterations in brain network organization that extend beyond the resolution of the overt clinical deficit. Moreover, the studies described above investigated brain network functioning and morphology in patients with presumed TIA within one month of the episode. Although these studies reported significant differences relative to healthy controls, they provide limited insight into the longitudinal evolution of these findings. Specifically, they cannot determine whether such alterations persist over time, resolve during recovery, or the extent to which brain structure and function may return toward normal levels.

To our knowledge, no previous study has investigated patients with TIA using fMRI during a motor paradigm specifically targeting their transient neurological deficits within the first 16 hours after symptom onset, followed by a longitudinal assessment of potential motor network reorganization. To address this gap, we included patients presenting to the emergency department with acute transient neurological symptoms clinically suggestive of a TIA, including transient motor or sensory symptoms, in the absence of an acute lesion on brain imaging. Importantly, the diagnosis of TIA could not be established solely on the basis of the initial clinical presentation and negative imaging findings, but relied on the subsequent clinical course and follow-up at 3 months. Accordingly, we explored longitudinal changes in motor-network activity and functional connectivity in patients presenting with a transient motor deficit mimicking an ischemic stroke, including both vascular and non-vascular etiologies. More specifically, we addressed three complementary questions. First, we investigated whether transient motor symptoms were associated with alterations in BOLD activity within the motor network during the acute phase (<16 h after symptom onset), and whether these alterations changed at 3-month follow-up in patients with TIA. We further examined whether acute BOLD alterations were related to the clinical lateralization of symptoms, with the expectation that abnormalities would be more pronounced in motor regions contralateral to the symptomatic limb. Second, we investigated task-related functional connectivity within the motor network using generalized psychophysiological interaction (gPPI) analysis. We asked whether patients with TIA and TIA mimics showed altered patterns of motor-network connectivity during the acute phase and, importantly, whether these connectivity patterns changed between the acute and follow-up assessments. Given the transient nature of the clinical symptoms and the absence of a visible structural lesion in many patients, we were particularly interested in whether longitudinal changes in functional connectivity could provide evidence of persistent or evolving functional alterations despite clinical resolution. Third, we explored whether longitudinal changes in motor-network activity and connectivity were shared across patients presenting with a TIA-like motor syndrome or differed according to the underlying etiology. This allowed us to determine whether the functional alterations associated with transient motor symptoms represented a common response to a transient neurological disturbance or whether distinct patterns were associated with the final clinical diagnosis. Overall, we aimed to characterize the functional signature of transient motor disturbances in the absence of an overt structural lesion and to determine the extent to which these changes were related to clinical symptom lateralization, longitudinal evolution, and etiological mechanism.

These preliminary findings suggest bilateral Bold decreased activation at the acute stage of TIA. This initial functional deficit tended to normalize over time, returning to baseline or near-baseline levels at 3 months, without the appearance of any lesion. This study has strong limitations primarily due to its sample size. Future works are needed to confirm the clinical validity of the biomarker and the pathophysiology it implies. However, this is a unique study presenting emergency fMRI in TIA, with detailed clinical and radiological definition of TIA which is fundamuntal in this pathology.

## Data Availability

All data produced in the present study are available upon reasonable request to the authors

## Supplementary material

**Supplementary Figure 1.**
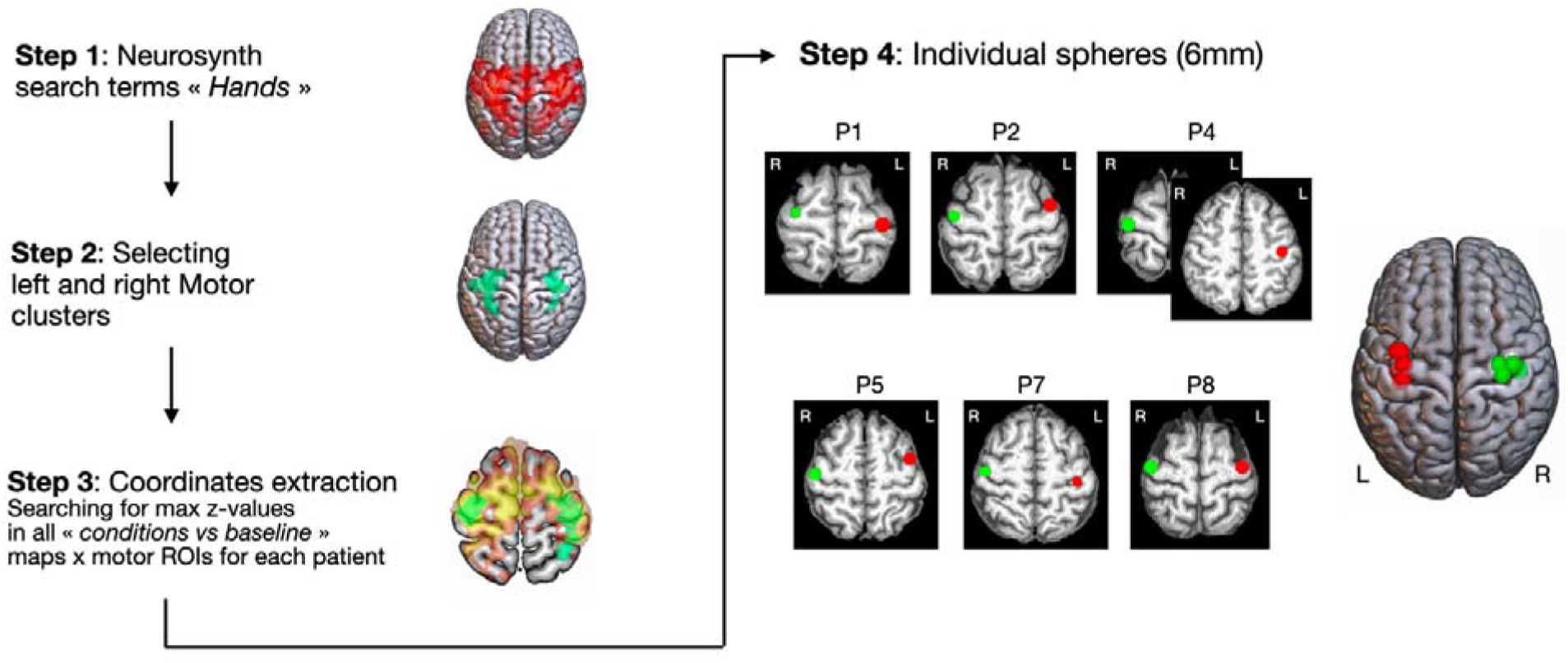
Representation of the different steps used to define the individual ROIs. Individual ROI spheres for the six patients are depicted on their individualized normalized T1-weighted maps, with the left side displayed according to radiological convention. Spheres corresponding to the left primary motor cortex (left M1) are shown in red, and those corresponding to the right primary motor cortex (right M1) in green. On the right, all spheres are projected onto a common MNI template provided by MRIcroGL.

**Supplementary Table 1.**
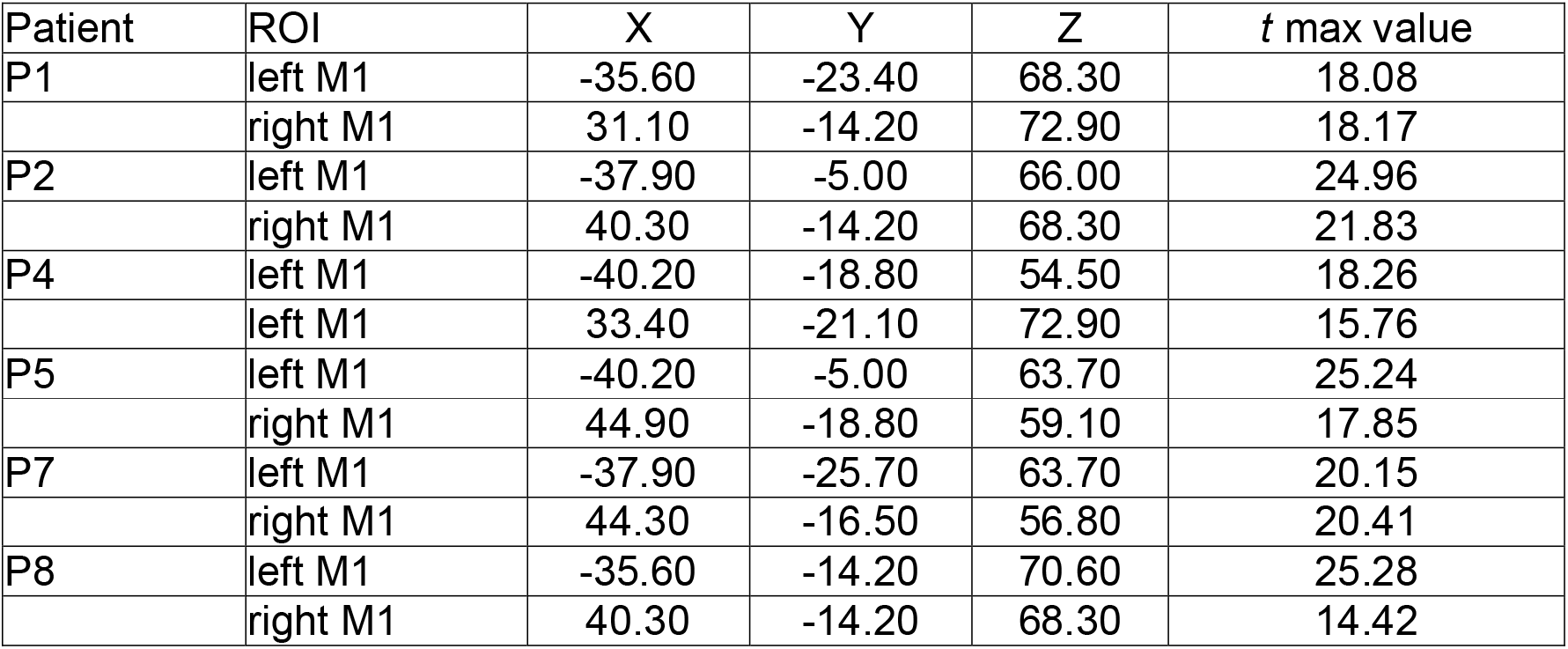
MNI coordinates of individual ROI sphere centers in the left and right primary motor cortex (M1) for each patient.

**Supplementary Table 2.**
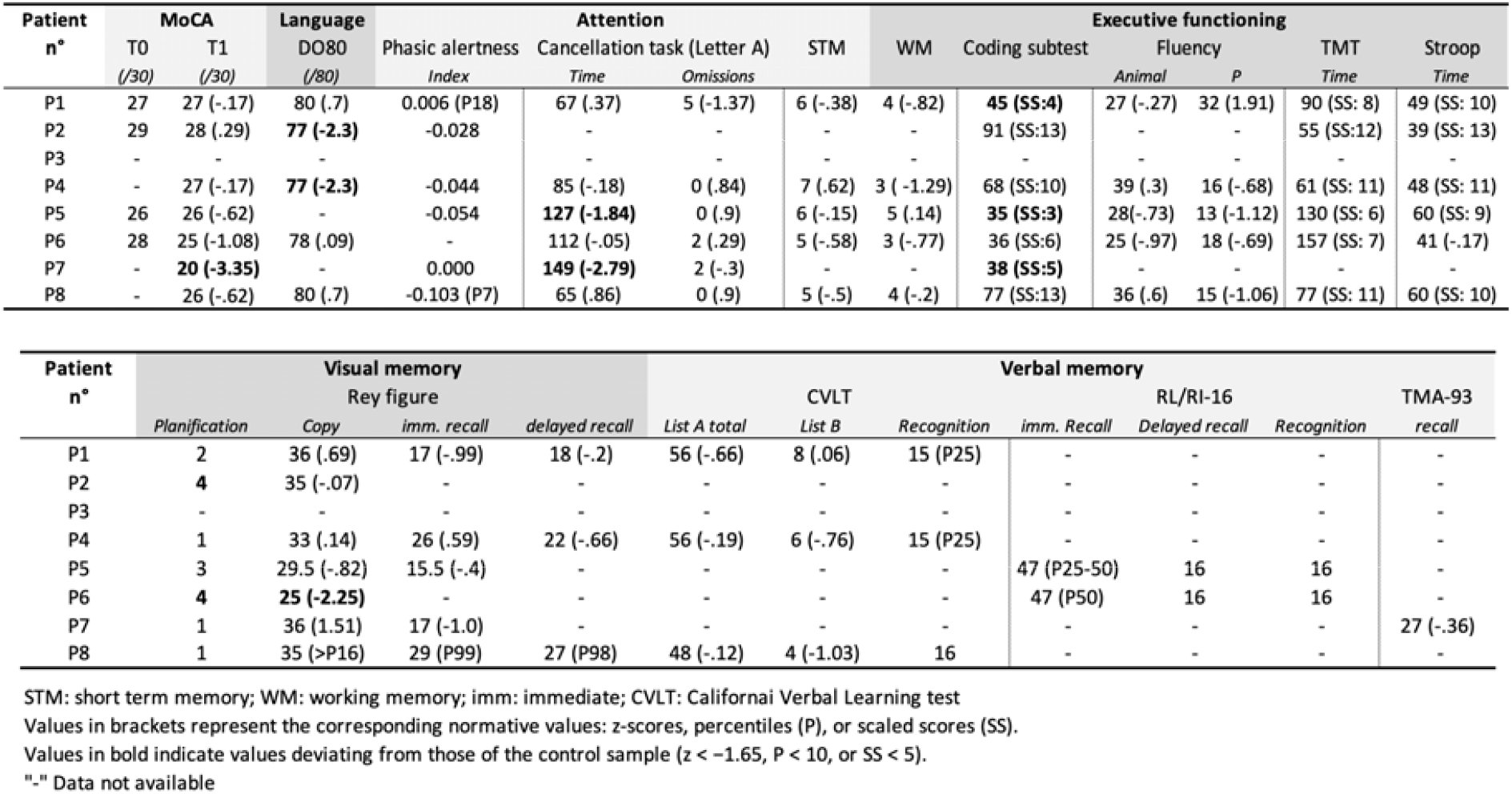
Neuropsychological data.

**Supplementary Table 3.** β values extracted from left M1 and right M1 for each patient during left- and right-hand movement conditions at T0 (acute stage) and T1 (chronic stage).

| Left M1 | T0 |  | T1 |  |
| --- | --- | --- | --- | --- |
|  | Right hand movement | Left hand movement | Right hand movement | Left hand movement |
| P1 | 5.22 | -0.08 | 0.15 | 0.13 |
| P2 | 3.29 | 0.50 | 4.00 | 0.93 |
| P4 | 0.67 | 0.75 | 3.31 | -0.04 |
| P5 | 0.84 | -0.05 | 6.54 | 3.14 |
| P7 | 3.68 | 1.20 | 3.96 | -0.56 |
| P8 | 1.18 | 0.28 | 2.36 | 1.2 |

| Right M1 | T0 |  | T1 |  |
| --- | --- | --- | --- | --- |
|  | Right hand movement | Left hand movement | Right hand movement | Left hand movement |
| P1 | 0.52 | 2.91 | 0.51 | 1.81 |
| P2 | -0.63 | 3.31 | 0.35 | 4.10 |
| P4 | -0.23 | 0.47 | -1.12 | 5.87 |
| P5 | 0.08 | -0.02 | -0.63 | 5.17 |
| P7 | 1.59 | 2.81 | 0.09 | 4.27 |
| P8 | -0.56 | 0.21 | -0.02 | 3.08 |

## Footnotes

1 The term “Hand” was selected, although alternative terms such as “Hands” and “Hand movements” yielded similar meta-analytical maps.

2 Patient 3 had no fMRI data available at T1, whereas Patient 6 had no fMRI data available at either T0 or T1.

